# D3MI: an efficient and powerful federated imputation method for bias reduction in the analysis of distributed incomplete data by accounting for within-site correlation and between-site heterogeneity

**DOI:** 10.1101/2025.05.08.25327224

**Authors:** Yi Lian, Xiaoqian Jiang, Qi Long

**Author notes:** Corresponding author: Yi Lian, Qi Long, Department of Biostatistics, Epidemiology and Informatics Perelman School of Medicine, University of Pennsylvania, 201 Blockley Hall, 423 Guardian Drive, Philadelphia, PA 19104.

## Abstract

**Objective:** Electronic health records (EHRs) collected from diverse healthcare institutions offer a rich and representative data source for clinical research. Federated learning enables analysis of these distributed data without sharing sensitive patient-level information, preserving privacy. However, missing data remain a major challenge and can introduce substantial bias if not properly addressed. Very few distributed imputation methods currently exist, and they fail to account for two critical aspects of EHR data: correlation within sites and variability across sites. We aim to fill this important methodological gap.

**Methods:** We propose Distributed Mixed Model-based Multiple Imputation (D3MI), a novel federated imputation method designed to reduce bias in distributed EHRs. D3MI integrates the strengths from federated learning techniques, statistical learning methods for correlated data, and multilevel imputation algorithms to explicitly account for both and within-site correlation and between-site heterogeneity using site-specific random effects. It preserves privacy by avoiding sharing raw data and features communication and computational efficiency.

**Results:** Through extensive simulation studies, we demonstrate that D3MI outperforms SOTA distributed imputation methods in both accuracy and consistency. We further demonstrate the use of D3MI in a real-world EHR case study involving incomplete and clustered data from participating hospitals in the Georgia Coverdell Acute Stroke Registry.

**Conclusion:** By explicitly modeling the complex structure of distributed EHR data, D3MI addresses key limitations of existing approaches. It provides a powerful and efficient solution for handling missing data in distributed and privacy-sensitive settings and enhances the rigor and reproducibility of collaborative clinical research.

## INTRODUCTION

Healthcare providers across the United States regularly gather electronic health records (EHRs), which have been extensively used for research. Collaborative research utilizing multiple EHRs from institutions in various geographic regions can create a larger and more representative sample of the U.S. population, leading to more reliable and generalizable research outcomes. This approach is particularly valuable for marginalized or underserved groups defined by sensitive attributes, as individual institutions may have highly imbalanced group compositions influenced by local demographics. However, these health data are generally stored and managed by the institutions who collect and own them, and individual patient data cannot be shared across sites due to regulations for the protection of personal health information. For instance, the Patient-Centered Scalable National Network for Effectiveness Research (pSCANNER) comprises data from 13 sites. These include the Veterans Affairs Health System, University of California Medical Centers, and University of Southern California, along with additional healthcare institutions contributing to the network.[1] The pSCANNER sites store, own and govern their own patient-level EHR data without a central repository that combines or merges data across sites.

A wide range of federated and distributed learning algorithms have been developed and employed to analyze decentralized healthcare data while maintaining patient confidentiality and data security.[2] However, for such multi-site data from diverse geographic locations, there exists heterogeneity between sources that needs to be accounted for. We use the Georgia Coverdell Acute Stroke Registry (GCASR) data as a motivating example.[3] The GCASR program aims to enhance the quality of care for acute stroke patients. One important indicator of this quality is the arrival-to-computed tomography (CT) time, as it significantly impacts patient survival.[4] Figure *1* shows the marginal distribution of the log arrival-to-CT time in the 15 largest hospitals in the GCASR data. While some differences in arrival-to-CT time can be explained by observed data, other variations may reflect different ‘baseline’ arrival-to-CT times across hospitals that may not be accounted for by the observed data. This also suggests that patients treated within the same hospital may be more similar to each other than to those from other hospitals, indicating potential within-hospital correlation. Failure to address the heterogeneity and correlation may lead to biased estimates and underestimation of the standard errors of the association.[5] Generalized linear mixed models (GLMM) can be used to tackle these challenges through the estimation of the hospital-specific random intercepts, while adjusting for factors (e.g., age, insurance coverage) that can potentially impact arrival-to-CT time. To our knowledge, three distributed GLMM algorithms have been proposed to analyze such distributed health data without sharing individual patient data across sites.[6-8] Detailed discussion on these methods can be found in the Supplementary Material.

**Figure 1.**
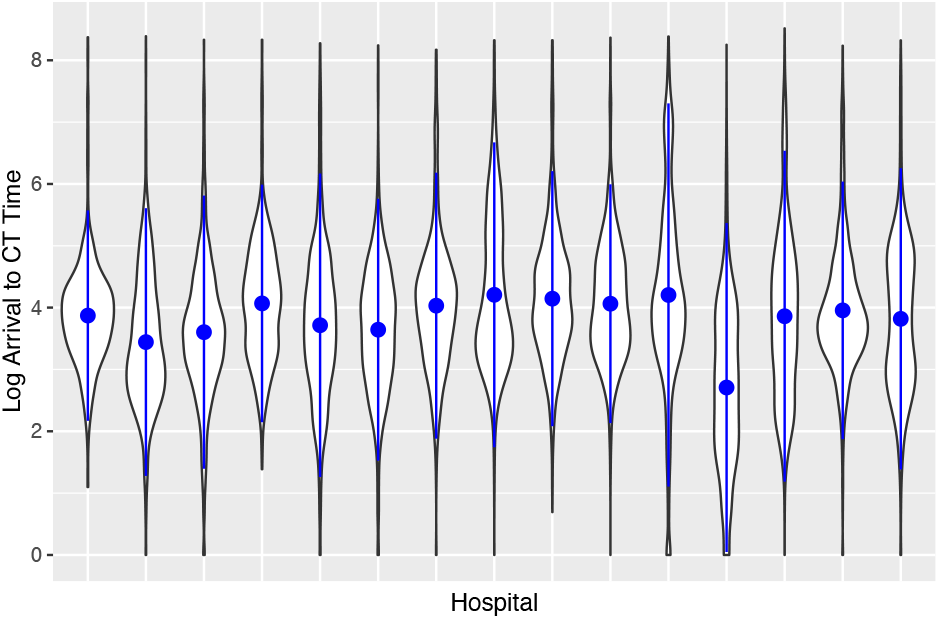
Distribution of log arrival-to-computed tomography time at the 15 largest hospitals in the Georgia Coverdell Acute Stroke Registry data

Furthermore, as incomplete observations at each site (e.g., hospital) is anticipated, missing data need to be addressed in the distributed analysis of decentralized healthcare data as well. Missing data problems can be more complex in distributed data as both the missing proportion and the distribution of the incomplete variables may vary. We again refer to the GCASR example to demonstrate the potential heterogeneity and correlation. A variable EMSnote, indicating whether the hospital received advance notice from the emergency medical services (EMS) about a stroke patient, is potentially associated with reduced arrival-to-CT time.[9] In order to verify the association (in **Real-world case study**), we first plot the distribution of the binary EMSnote in the 15 largest hospitals in Figure 2.

**Figure 2.**
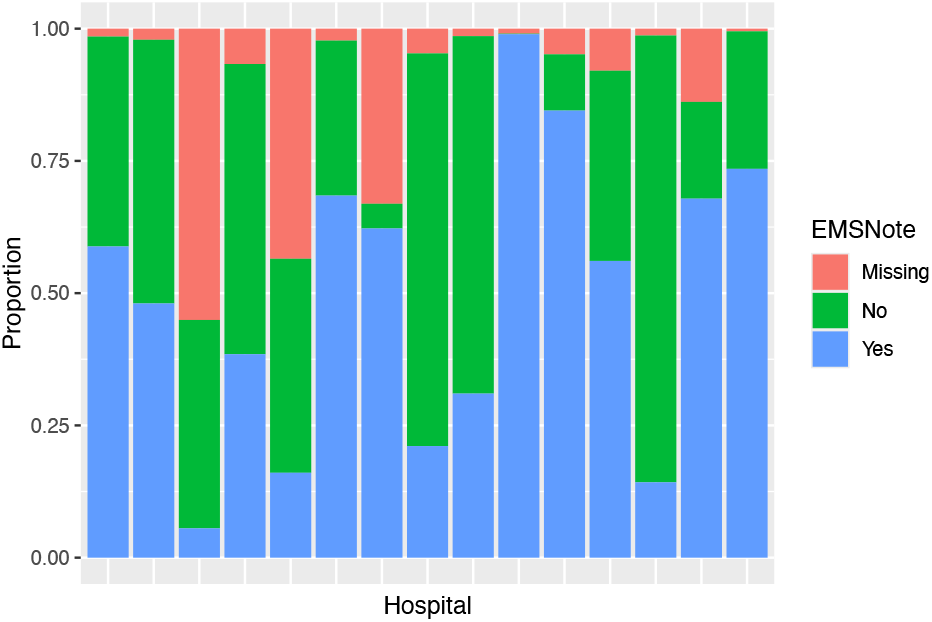
Distribution of the binary variable indicating whether the hospital received advance notice from emergency medical services (EMSnote) and the missing proportions at the 15 largest hospitals in the Georgia Coverdell Acute Stroke Registry data

The missing proportion of this variable range from below 1% to over 55%. Additionally, based on the observed data, the proportion of patients receiving EMSnote varies widely across hospitals, ranging from as low as 12% to nearly 100%. These stark differences highlight substantial variability in distributed healthcare data, potentially driven by differences in care practices and data collection protocols across sites, which likely contribute to greater similarity among patients within the same hospital. Throughout the paper, we assume that the missingness in the incomplete variables are dependent on the fully observed ones, e.g. missing at random (MAR).

In this paper, we propose a Distributed Mixed Model-based Multiple Imputation (D3MI) method to address missing data problems in the analysis of decentralized healthcare data featuring between-site heterogeneity and within-site correlation. Leveraging privacy-preserving distributed GLMM algorithms, the D3MI method does not require sharing of individual patient data across different sites. With **Simulation** studies, we show that D3MI outperforms current SOTA distributed imputation algorithms that ignore heterogeneity and correlation in distributed data. With a single one-way communication of summary statistics per missing variable, D3MI can effectively achieve satisfactory and consistent imputation performance. In **Real-world case study**, D3MI generates significantly different model estimates than SOTA imputation methods.

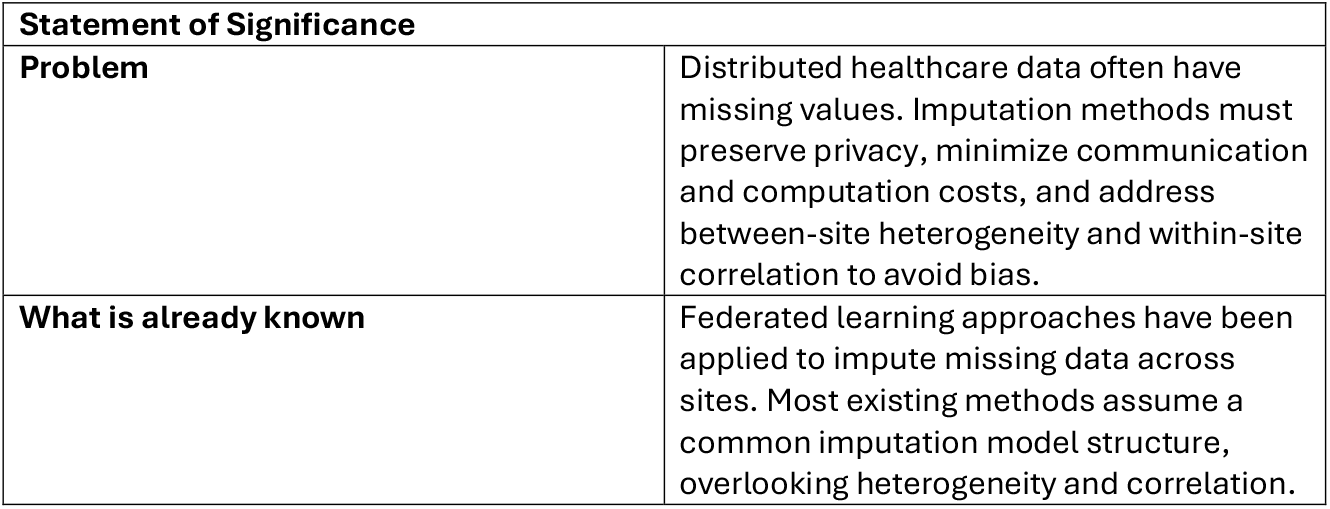
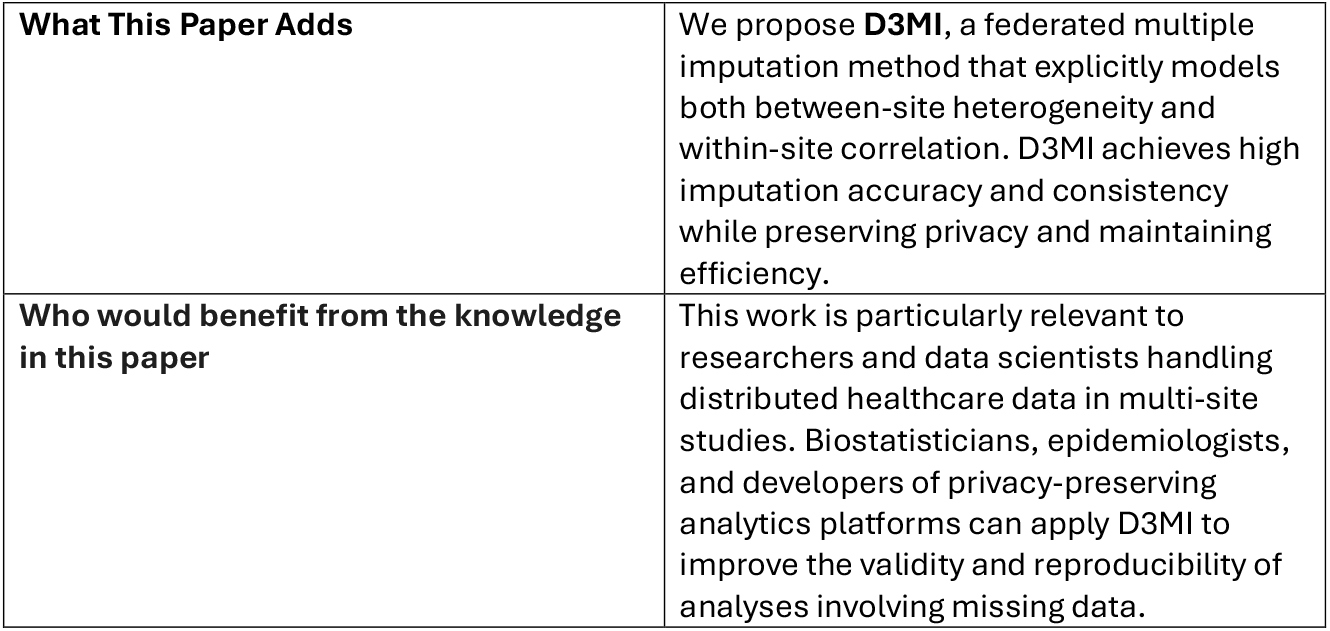

| Statement of Significance |  |
| --- | --- |
| **Problem** | Distributed healthcare data often have missing values. Imputation methods must preserve privacy, minimize communication and computation costs, and address between-site heterogeneity and within-site correlation to avoid bias. |
| **What is already known** | Federated learning approaches have been applied to impute missing data across sites. Most existing methods assume a common imputation model structure, overlooking heterogeneity and correlation. |
| **What This Paper Adds** | We propose **D3MI**, a federated multiple imputation method that explicitly models both between-site heterogeneity and within-site correlation. D3MI achieves high imputation accuracy and consistency while preserving privacy and maintaining efficiency. |
| **Who would benefit from the knowledge in this paper** | This work is particularly relevant to researchers and data scientists handling distributed healthcare data in multi-site studies. Biostatisticians, epidemiologists, and developers of privacy-preserving analytics platforms can apply D3MI to improve the validity and reproducibility of analyses involving missing data. |

## RELATED WORK

Multilevel imputation methods have been proposed to address missing data problems in data collected from diverse sites that are correlated within each site.[10-17] Some of these methods involve mixed models, which account for the between-site heterogeneity and within-site correlations in the covariate with missingness.[15, 16] However, none of these methods are developed for distributed learning settings. In addition, there have been limited work on distributed/federated multiple imputation methodology.[18-21] Current state-of-the-art (SOTA) imputation methods, such as the averaged mixture (AVGM)-based algorithm in Chang et al. (2020) [18] and the FedIMPUTE-Pooled algorithm in Li et al. (2025) [21], estimate global imputation models by computing a weighted average of local model estimates across sites. Chang et al. also proposes an alternative method based on the communication-efficient surrogate likelihood (CSL) algorithm [22]. Similarly, Li et al. also introduces FedIMPUTE-DAC, which builds on the divide-and-conquer (DAC) framework.[23] Both the CSL- and DAC-based approaches rely on obtaining initial imputation model estimates from a central site and updating them using pooled first- and/or second-order information from all sites. The performance of these methods relies on a central site with relatively large sample size or low missing rate. For example, the software implementation of FedIMPUTE-DAC requires a fully observed central site, therefore it is not applicable to the setting of our interest. Finally, none of these distributed imputation methods were designed to account for multilevel data featuring between-site heterogeneity and within-site correlation as they use a unified imputation model for all sites. Our work seeks to address this significant methodological gap.

## METHODS

Suppose we have data from *i* = 1, … , *L* sites and each site *i* has *j* = 1, … , *n*_*i*_ observations. For the *j*-th observation from the *i*-th site, we denote the outcome variable by *y*_*ij*_ and the set of *p* covariates by *X*_*ij*_. The GLMM with a random intercept has the form

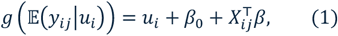

where *g*(⋅) is the link function, e.g. logit function for binary outcomes and identity link function for continuous outcomes, 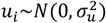 is the site-specific random intercept, and (*β*_0_, *β*) are the fixed effects. Note that the GLMM uses the same set of fixed effect parameters to estimate the association between the covariates and the outcome, and the heterogeneity between the sites can be quantified by 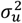. The common fixed effect terms allow for information sharing, which is particularly beneficial for sites and groups with small sample sizes. Further, the random effect *u*_*i*_ in (**Error! Reference source not found**.) for each site *i* can be subsequently estimated if it is of interest to the researcher.

The GLMM-based D3MI method has three main components like other multiple imputation algorithms, which include imputation, analysis and pooling. To preserve data privacy, the first two components are performed in a distributed manner following previous work.[18, 19] These components of the D3MI are adapted from existing research (discussed in the Supplementary Material) that are suitable for the distributed multiple imputation considering computational and communication efficiency.

### Distributed imputation

As discussed in the motivating example in the **INTRODUCTION** section, the incomplete covariates may also be subject to within-site correlations and between-site heterogeneity. Therefore, we propose to use distributed GLMM-based multiple imputation to account for the potential heterogeneity and correlation. Specifically, a GLMM model like 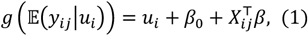 is fitted for each incomplete covariate,

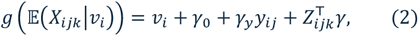

where *k* is the index of the covariate that has missing values, *X*_*ijk*_ is the value of the *k*-th covariate of the *j*-th patient at the *i*-th site and *Z*_*ijk*_ denotes the vector of covariates included in the imputation model, 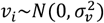 is the random intercept of the *i*-th site and ***γ*** = (*γ*_0_, *γ*_*y*_, *γ*) are the fixed effects. Like conventional multiple imputation for MAR data based on LMs and GLMs, the covariates *Z*_*ijk*_ chosen to impute the missing covariate *X*_*ijk*_ should be associated with *X*_*ijk*_ or the occurrence of its missingness. We adopt the distributed PQL (dPQL) approach to fit these GLMM imputation models in a privacy-preserving manner.[8] This approach features high computational and communication efficiency (See Supplementary Material for description and comparison with alternatives). For Gaussian GLMM with the identity link for continuous variable, both the dPQL and its original PQL can converge to the estimates in one iteration.[8] For binary GLMM with the logit link, a few iterations or even one iteration will suffice for the purpose of imputation.

Using the dPQL, we acquire the GLMM estimates 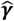and 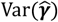 (as well as the residual variance for Gaussian GLMM with identity link) and the predicted 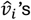. We then adopt the approximate Bayesian approach for multilevel imputation (See **Supplementary Material**) to generate random draws of ***γ*** and 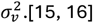 Finally, with the randomly drawn parameters, missing values in the covariate are randomly generated for a total of *M* times. When multiple covariates are missing, we incorporate our distributed imputation method within the multivariate imputation with chained equation (MICE) framework to iteratively impute each covariate based on the other covariates.[24]

### Distributed analysis

With the construction of multiple complete datasets through distributed multiple imputation, the next step is to perform distributed analysis on the imputed datasets. As discussed previously, the analysis model of choice is also GLMM, to account for the heterogeneity and correlation. The model has been given in Equation 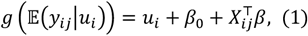. In the GCASR example, such GLMM model estimates the association between arrival-to-CT time and EMSnote, adjusting for covariates and accounting for the heterogeneity between sites through the random intercept. In addition, this GLMM assumes that the observations are independent conditional on the random effects, accounting for potential between-site correlations, e.g., patients that go to the same hospital may be correlated. Again, we employ the dPQL approach to estimate the model coefficients 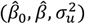.[8]

### Pooling

In the last step, estimates from the distributed analyses of the imputed datasets are pooled to generate the results following Rubin’s rule. For both fixed effects (*β*_0_, *β*) and the variance of the random intercept 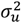, the final pooled estimates can be computed by taking the mean of the estimates acquired in the analyses of imputed datasets. The variance-covariance matrix of the fixed effects (to generate the confidence intervals) can also be computed according to the Rubin’s rule,[25] which has been established in previous work in centralized multilevel imputation and implemented in the `mice` package.[15, 16, 24] Details can be found in the **Supplementary Material**.

We summarize the D3MI algorithm below:

1. For each covariate with missingness
  a. Fit a GLMM with site-specific random intercept, with the covariate with missingness as the dependent variable. The predictors include other covariates (observed or previously imputed) and the outcome variable of the final analysis.
  b. Estimate the GLMM parameters (fixed effects with variance, variance of the random intercept, and the site-specific random intercept) using dPQL approach.[8]
  c. Generate random draws of the fixed effects, random intercept (and additionally residual standard error for continuous missing variable) based on the parameters estimated in Step 1b.[16]
  d. Impute missing values in the covariate based on the random parameters generated in Step 1c for a total of *M* times.[16]
2. Fit a distributed GLMM for each imputed datasets with site-specific random intercept to estimate the association between the outcome variable of the analysis model and the imputed covariates using dPQL.
3. Pool the *M* sets of estimated model parameters from Step 2 following Rubin’s rule.[25]

The D3MI algorithm by default takes one iteration in Steps 1a and 2 to minimize computational and communication costs during both distributed imputation and analysis. In this sense, the D3MI can be regarded as a communication-efficient distributed algorithm.

## RESULTS

### Simulation

We perform extensive simulation studies to examine the performance of D3MI. We generate a continuous variable *x*_*i*3_ ∼ *N*(0, 0.5^2^) for the *i*-th site, a continuous variable

*x*_*i*2_∼*N*(*u*_*i*2_ + 0.1*x*_3_, 0.1^2^) with random intercept *u*_*i*2_ ∼ *N*(0,0.1^2^) for *i* = 1, … , *L*, and a binary variable *x*_*i*1_ ∼ *Binom*(1*/*(1 + exp(−*u*_*i*1_ − 0.1*x*_*i*2_ − 0.1*x*_*i*3_))) with random intercept *u*_*i*1_ ∼ *N*(0, 0.1^2^). The outcome *y* is generated using *y*_*i*_ ∼ *N*(−0.5 + 0.5*x*_*i*1_ + 0.5*x*_*i*2_ + 0.1*x*_*i*3_ + *u*_*iy*_, 0.5^2^) with random intercept 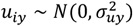. We consider three scenarios, a) *x*_1_ is incomplete; b) *x*_2_ is incomplete; and c) *x*_1_ and *x*_2_ are both incomplete. In all three scenarios, we assume the missing mechanism is MAR. In each scenario, we consider a total sample size of either *N* = **600** or *N* = **1200**, evenly distributed in **5** or **10** sites and two levels of between-site heterogeneity (**strong**: 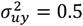 and **weak**: 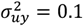). The missing probability for *x*_1_ is generated randomly based on a logistic model involving an intercept, *x*_2_, *x*_3_ and *y* following an assumed MAR mechanism, and the same applies to *x*_2_. The intercept is adjusted such that, in all scenarios, the missing proportion in *x*_1_ and/or *x*_2_ across sites is roughly 15-20%.

We compare the performance of D3MI with two distributed imputation methods as competitors and four baseline approaches as benchmarks. The competitors are two SOTA computation-efficient and privacy-preserving distributed multiple imputation algorithms based on linear models (LM) and generalized LMs (GLM) without considering clustering of the distributed data.[18] The distributed imputation is achieved through the **AVGM** and the **CSL** algorithms.[22, 26] For fair comparison, the distributed analyses following AVGM- and CSL-based imputation are performed using dPQL[8] and the results are pooled following Rubin’s rule.[25] We denote the non-distributed version of D3MI, referred to as **PQL**, which represents the optimal performance achievable by D3MI—due to its lossless property[8]— when iterated to convergence. It also serves as a bridge for comparison with the next competitor, **lme4**, which refers to the `lme4` package in the R software. The package is widely considered to generate the most accurate GLMM estimates, and lme4-based multilevel multiple imputation[15, 16] has been implemented in the go-to R package `mice` (details in the Supplementary Material).[27] Therefore, the hypothetical centralized lme4’s performance can be considered as the gold standard that distributed GLMM-based multiple imputation methods should aim to approach. Finally, we perform a hypothetical complete data (**CD**) analysis (that uses all the generated data including the missing values) and a complete case (**CC**) analysis (that uses all the observed data) as benchmarks. No imputation is performed in these two benchmarks, and we perform GLMM-based analyses directly.

With all the aforementioned methods, *M* = 5 imputations are performed with 10 iterations between imputation, and the same procedure is replicated on 1000 Monte Carlo simulated datasets.[16] Following previous work,[18, 19] we compare the relative **Bias**: 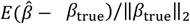 , standard deviation of the 1000 estimates (**SD**): 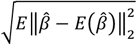 and root mean squared error (**rMSE**): 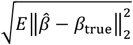 . We summarize the results for evenly distributed sites in Table *1*, Table *2* and Table *3*.

**Table 1.** Simulation scenario a) one binary variable has missingness. Mixed model-based imputation methods are in **bold**, distributed imputation methods are in *italics*. Benchmark methods are in shaded cells, including two analyses without imputation and two centralized imputation methods.

| Heterogeneity | No. of Sites | Method | $N = 600$ | | | $N = 1200$ | | |
| --- | --- | --- | --- | --- | --- | --- | --- | --- |
|  |  |  | Bias | SD | rMSE | Bias | SD | rMSE |
| Weak | 5 | CD | 0.002 | 0.214 | 0.213 | 0.002 | 0.157 | 0.157 |
|  |  | CC | 0.093 | 0.229 | 0.243 | 0.081 | 0.171 | 0.185 |
|  |  | **lme4** | 0.003 | 0.218 | 0.218 | 0.005 | 0.161 | 0.161 |
|  |  | **PQL** | 0.003 | 0.220 | 0.220 | 0.006 | 0.163 | 0.163 |
|  |  | *AVGM* | 0.004 | 0.267 | 0.267 | 0.016 | 0.241 | 0.241 |
|  |  | CSL | 0.010 | 0.288 | 0.288 | 0.017 | 0.259 | 0.259 |
|  |  | **D3MI** | 0.009 | 0.219 | 0.220 | 0.021 | 0.162 | 0.163 |
|  | 10 | CD | 0.002 | 0.200 | 0.200 | 0.008 | 0.148 | 0.148 |
|  |  | CC | 0.088 | 0.223 | 0.235 | 0.096 | 0.161 | 0.181 |
|  |  | **lme4** | 0.006 | 0.206 | 0.206 | 0.007 | 0.151 | 0.151 |
|  |  | **PQL** | 0.007 | 0.208 | 0.208 | 0.007 | 0.152 | 0.152 |
|  |  | *AVGM* | 0.016 | 0.241 | 0.241 | 0.013 | 0.193 | 0.193 |
|  |  | CSL | 0.017 | 0.259 | 0.259 | 0.006 | 0.213 | 0.213 |
|  |  | **D3MI** | 0.022 | 0.207 | 0.208 | 0.016 | 0.152 | 0.152 |
| Strong | 5 | CD | 0.007 | 0.312 | 0.312 | 0.003 | 0.272 | 0.272 |
|  |  | CC | 0.093 | 0.326 | 0.336 | 0.086 | 0.282 | 0.292 |
|  |  | **lme4** | 0.016 | 0.316 | 0.316 | 0.014 | 0.275 | 0.275 |
|  |  | **PQL** | 0.016 | 0.316 | 0.316 | 0.014 | 0.275 | 0.275 |
|  |  | *AVGM* | 0.065 | 1.023 | 1.019 | 0.090 | 1.041 | 1.038 |
|  |  | CSL | 0.099 | 1.103 | 1.103 | 0.122 | 1.126 | 1.127 |
|  |  | **D3MI** | 0.026 | 0.316 | 0.316 | 0.025 | 0.275 | 0.276 |
|  | 10 | CD | 0.009 | 0.265 | 0.265 | 0.015 | 0.219 | 0.219 |
|  |  | CC | 0.092 | 0.286 | 0.297 | 0.100 | 0.229 | 0.245 |
|  |  | **lme4** | 0.020 | 0.271 | 0.271 | 0.018 | 0.223 | 0.224 |
|  |  | **PQL** | 0.020 | 0.271 | 0.271 | 0.018 | 0.224 | 0.224 |
|  |  | *AVGM* | 0.082 | 0.813 | 0.808 | 0.086 | 0.758 | 0.753 |
|  |  | CSL | 0.105 | 0.880 | 0.878 | 0.088 | 0.823 | 0.819 |
|  |  | **D3MI** | 0.032 | 0.270 | 0.272 | 0.024 | 0.223 | 0.224 |

**Table 2.** Simulation scenario b) one continuous variable has missingness. Mixed model-based imputation methods are in **bold**, distributed imputation methods are in *italics*. Benchmark methods are in shaded cells, including two analyses without imputation and two centralized imputation methods.

| Heterogeneity | No. of Sites | Method | N = 600 |  |  | N = 1200 |  |  |
| --- | --- | --- | --- | --- | --- | --- | --- | --- |
|  |  |  | Bias | SD | rMSE | Bias | SD | rMSE |
| Weak | 5 | CD | 0.002 | 0.214 | 0.214 | 0.002 | 0.157 | 0.157 |
|  |  | CC | 0.083 | 0.230 | 0.241 | 0.080 | 0.171 | 0.185 |
|  |  | **lme4** | 0.003 | 0.239 | 0.239 | 0.002 | 0.177 | 0.177 |
|  |  | **PQL** | 0.003 | 0.239 | 0.239 | 0.002 | 0.177 | 0.177 |
|  |  | *AVGM* | 0.013 | 0.255 | 0.254 | 0.004 | 0.228 | 0.226 |
|  |  | *CSL* | 0.023 | 0.315 | 0.314 | 0.043 | 0.292 | 0.294 |
|  |  | **D3MI** | 0.003 | 0.238 | 0.238 | 0.002 | 0.177 | 0.177 |
|  | 10 | CD | 0.002 | 0.200 | 0.200 | 0.008 | 0.148 | 0.148 |
|  |  | CC | 0.083 | 0.218 | 0.229 | 0.084 | 0.161 | 0.177 |
|  |  | **lme4** | 0.005 | 0.223 | 0.223 | 0.011 | 0.165 | 0.165 |
|  |  | **PQL** | 0.006 | 0.223 | 0.223 | 0.011 | 0.165 | 0.165 |
|  |  | *AVGM* | 0.007 | 0.237 | 0.236 | 0.015 | 0.189 | 0.187 |
|  |  | *CSL* | 0.036 | 0.284 | 0.286 | 0.031 | 0.240 | 0.240 |
|  |  | **D3MI** | 0.006 | 0.223 | 0.223 | 0.011 | 0.165 | 0.165 |
| Strong | 5 | CD | 0.007 | 0.312 | 0.312 | 0.003 | 0.272 | 0.272 |
|  |  | CC | 0.084 | 0.330 | 0.338 | 0.080 | 0.283 | 0.291 |
|  |  | **lme4** | 0.009 | 0.334 | 0.334 | 0.003 | 0.286 | 0.286 |
|  |  | **PQL** | 0.008 | 0.335 | 0.335 | 0.003 | 0.287 | 0.287 |
|  |  | *AVGM* | 0.055 | 0.889 | 0.883 | 0.120 | 0.919 | 0.920 |
|  |  | *CSL* | 0.115 | 1.113 | 1.116 | 0.031 | 1.120 | 1.117 |
|  |  | **D3MI** | 0.009 | 0.335 | 0.335 | 0.003 | 0.286 | 0.287 |
|  | 10 | CD | 0.009 | 0.265 | 0.265 | 0.015 | 0.219 | 0.219 |
|  |  | CC | 0.083 | 0.285 | 0.294 | 0.087 | 0.229 | 0.241 |
|  |  | **lme4** | 0.011 | 0.292 | 0.292 | 0.014 | 0.234 | 0.234 |
|  |  | **PQL** | 0.010 | 0.292 | 0.293 | 0.014 | 0.234 | 0.235 |
|  |  | *AVGM* | 0.118 | 0.919 | 0.920 | 0.050 | 0.673 | 0.668 |
|  |  | *CSL* | 0.031 | 1.121 | 1.118 | 0.096 | 0.929 | 0.929 |
|  |  | **D3MI** | 0.011 | 0.292 | 0.292 | 0.014 | 0.234 | 0.235 |

In Table *1*, we summarize the results for simulation scenario a). Across the board, we can see that D3MI generates similar biases to AVGM and CSL when the between-site heterogeneity is weak, with much lower variations measured by SD and rMSE. When the heterogeneity is strong, D3MI clearly outperforms both AVGM and CSL. The very high bias, SD and rMSE generated by AVGM and CSL suggest that the ignoring the clustering effect can seriously harm the accuracy and consistency of the imputation. The very unstable performance of CSL could be explained by the fact that it heavily depends on a central site, where initial estimates are generated to construct the surrogate likelihood.[18, 22] Naturally, this method cannot be readily applied when there exist strong heterogeneity between the sites because the same surrogate likelihood may not be valid across all sites. On the other hand, D3MI (with one iteration) yields larger bias but similar SD and rMSE comparing to the hypothetical centralized lme4 and PQL. In this simulation setting, the suboptimal performance of D3MI compared to the benchmarks suggests that we can let D3MI run for a few more iterations (with a few more rounds of communication) to improve the imputation performance. As the number of iterations increases, we can expect the bias to decrease to the level generated by PQL. Additionally, the performance of PQL is consistently comparable to that of lme4. This suggests that, even though PQL may not be as accurate as the lme4 for estimating the same GLMM, the difference in imputation performance can is negligible.

In Table *2*, we see a similar overall pattern, which unfolds as follows. First, D3MI consistently outperforms the superior algorithm, whether it is AVGM or CSL, by a small margin under weak heterogeneity and by a significant margin under strong heterogeneity. Again, AVGM and CSL generate significantly higher SD and rMSE, particularly when the heterogeneity is strong, suggesting highly inconsistent imputation performance. GLMM-based imputation provides significant improvements, in terms of both accuracy and consistency, over (G)LM-based imputation methods that do not account for between-site heterogeneity. Next, we see that the performance of D3MI is almost identical to PQL and lme4 in imputing continuous variables by a Gaussian GLMM with identity link. This suggests that one iteration of computational and communication is sufficient for D3MI to achieve the best possible performance GLMM-base imputation can achieve, i.e., with the gold-standard GLMM solver under the centralized setting. On the other hand, if we compare the performance of the same methods when the total sample size is different (600 vs. 1200), we can see that all GLMM-based methods enjoy improved performance for larger sample size (lower bias, SD and rMSE). If the total sample size is fixed but the number of sites increases from 5 to 10, bias can increase slightly but the SD and rMSE both decreases. This may be due to the requirement of GLMMs for a sufficient number of sites to produce reliable estimates for the random intercept.

In Table 3 we illustrate scenario c) with a general pattern of missing data, where multiple covariates, both continuous and binary, exhibit missingness. Like the previous two scenarios, D3MI have better performance than AVGM and CSL, particularly when the between-site heterogeneity is large. While CSL may produce low biases, it exhibits significant variability, as indicated by the largest standard deviation (SD) and root mean square error (rMSE) compared to other methods. As we have discussed in simulation scenarios a) and b), (G)LM based AVGM and CSL cannot be reliably applied to impute heterogeneous data. In addition, the performance of these (G)LM-based imputation deteriorates as the number of heterogeneous sites increases. In contrast, lme4, PQL and D3MI show better performance (lower bias, SD and rMSE) when the number of sites increases, thereby improving the estimation the variance of the random intercept. Finally, we can see that D3MI’s performance is very close to lme4 and PQL across the board, with slightly larger bias, as well as SD and rMSE. The results suggest that when there is a mixture of continuous and binary missing variables, the communication-efficient D3MI algorithm can achieve performance comparable to that of the hypothetical centralized imputation using the ‘gold standard’ GLMM solver.

**Table 3.**
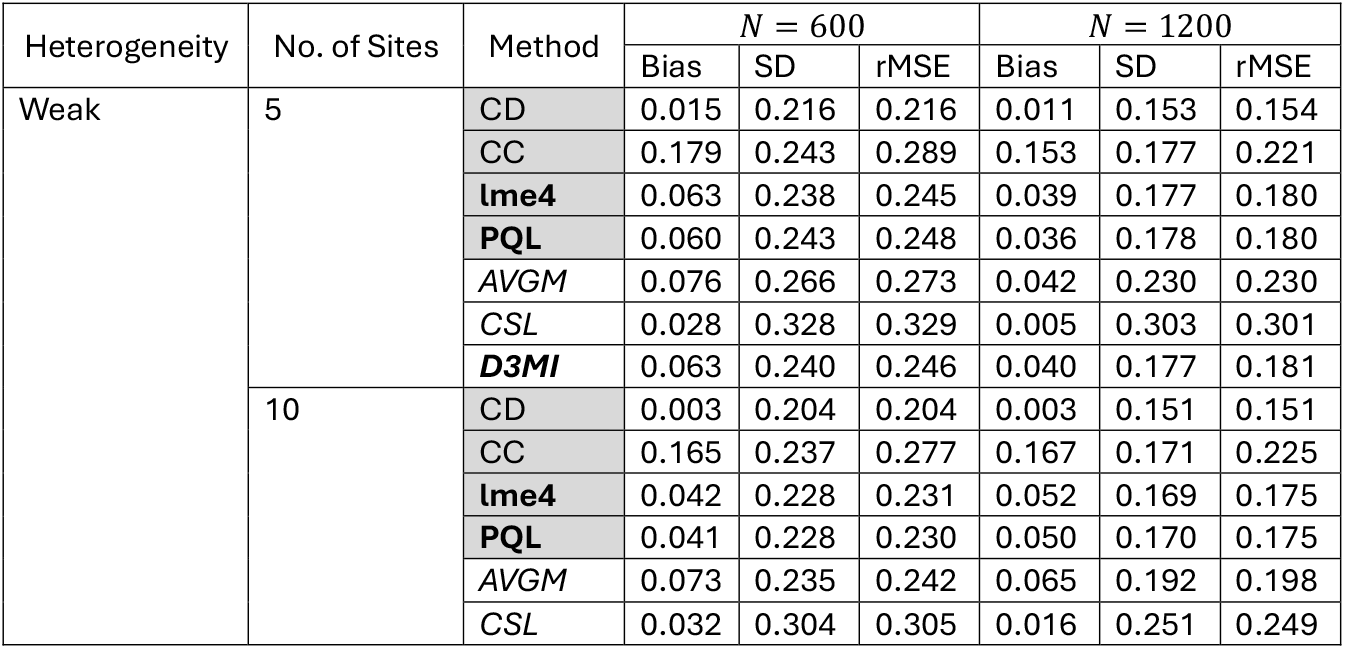
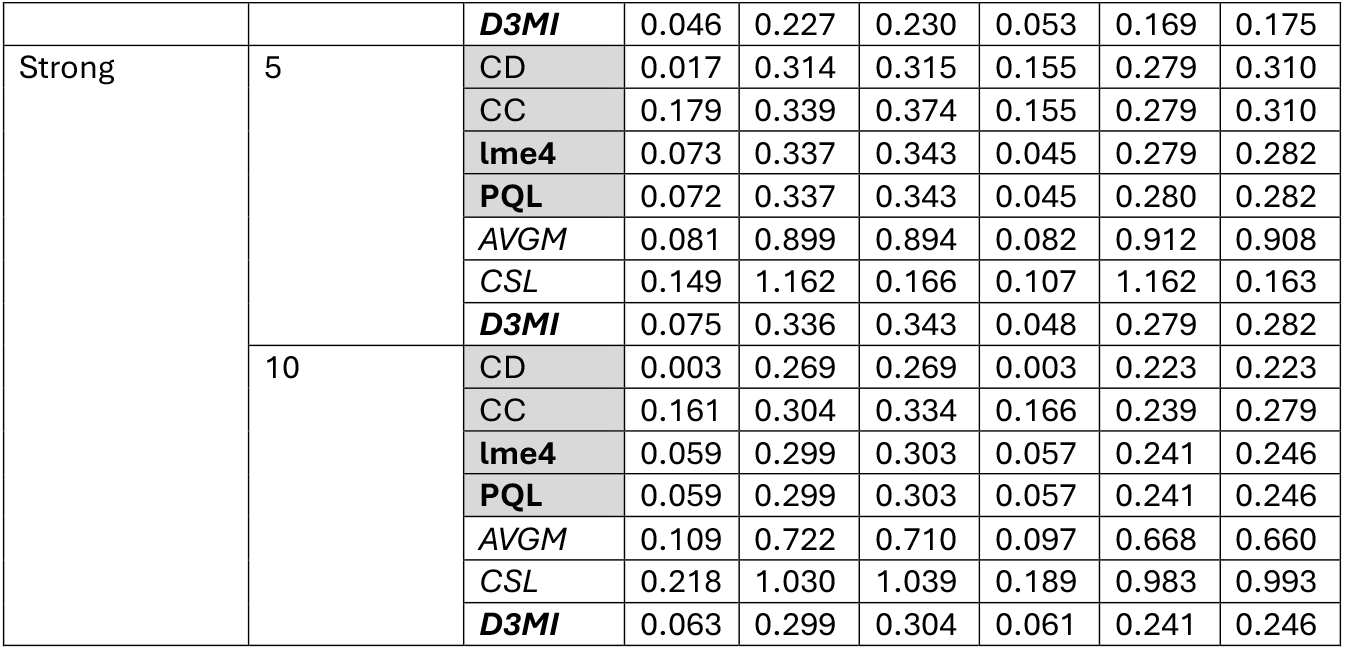
Simulation scenario c) both the binary variable and the continuous variable have missingness. Mixed model-based imputation methods are in **bold**, distributed imputation methods are in *italics*. Benchmark methods are in shaded cells, including two analyses without imputation and two centralized imputation methods.

### Real-world case study

We apply the D3MI method to a real-world case study using the GCASR data. As noted in the **INTRODUCTION** section, the GCASR provides information aimed at enhancing acute stroke care from various participating hospitals. In this case study, we aim to estimate the association between arrival-to-CT time and EMSnote, adjusting for potential confounders and between-hospital heterogeneity. The main aim of this case study is to explore how different imputation algorithms can affect the final estimates. To assemble the analysis cohort, we excluded hospitals with less than 10 patients and patients with arrival-to-CT time above five thousand minutes (deemed outliers based on the inspections of the distribution of arrival-to-CT time). The cohort consists of 36422 patients from 67 participating hospitals. We included several potential confounders defined a priori in the analysis including health insurance status (**Insured** vs. uninsured), race (**White** vs. other), **Gender**, arrived on a **Weekend** vs. a weekday, arrived during the **Day** vs. nighttime, and whether there has been documentation on patient and/or caregiver receiving education and/or resource materials regarding how to activate EMS for stroke (**EMSeduc**). There is missing data in **Insured** (1%), **Day** (1%), **EMSnote** (8%) and **EMSeduc** (14%). Finally, the outcome variable arrival-to-CT time is log transformed due to its significant right-skewedness.

We apply the same competitor and benchmark methods used in the **Simulation** section, including AVGM, CSL, CC, lme4 and PQL. We have access to aggregated data therefore the competing algorithms and the baseline algorithms can be applied for comparison. For the D3MI, we treat the hospital-specific datasets as if they cannot be shared. We also use complete case analysis with a random-intercept linear mixed model as a benchmark. For AVGM and CSL, we fit a distributed GLMM for the analysis after the imputation using the dPQL algorithm.[8] The MICE procedure is applied iteratively to impute the four incomplete covariates.[24] We summarize the estimates in Figure *3*. For all nine covariates (including the intercept), D3MI (with one iteration in imputation models and the analysis model) generates results very similar to those obtained through hypothetical centralized imputation using a gold standard GLMM solver. This observation agrees with the simulation results and suggests that the D3MI can generate highly satisfactory imputation results at minimal computational and communication cost. On the other hand, AVGM and CSL generate similar estimates that are substantially different from both GLMM-based imputation and the CC benchmark, which could be a result of not addressing the between-hospital heterogeneity and information loss during distributed imputation. Overall, after adjusting for the potential confounders, EMSnote is associated with a decrease of 0.24 (95% confidence interval [CI]: 0.20 to 0.27) in the log arrival-to-CT time based on the distributed imputation and analysis of the GCASR data using distributed GLMM, significantly different from those generated by SOTA distributed GLM-based multiple imputation (AVGM: 0.17, 95% CI: [0.14, 0.20]; CSL: 0.17, 95% CI: [0.14, 0.19]).

**Figure 3.**
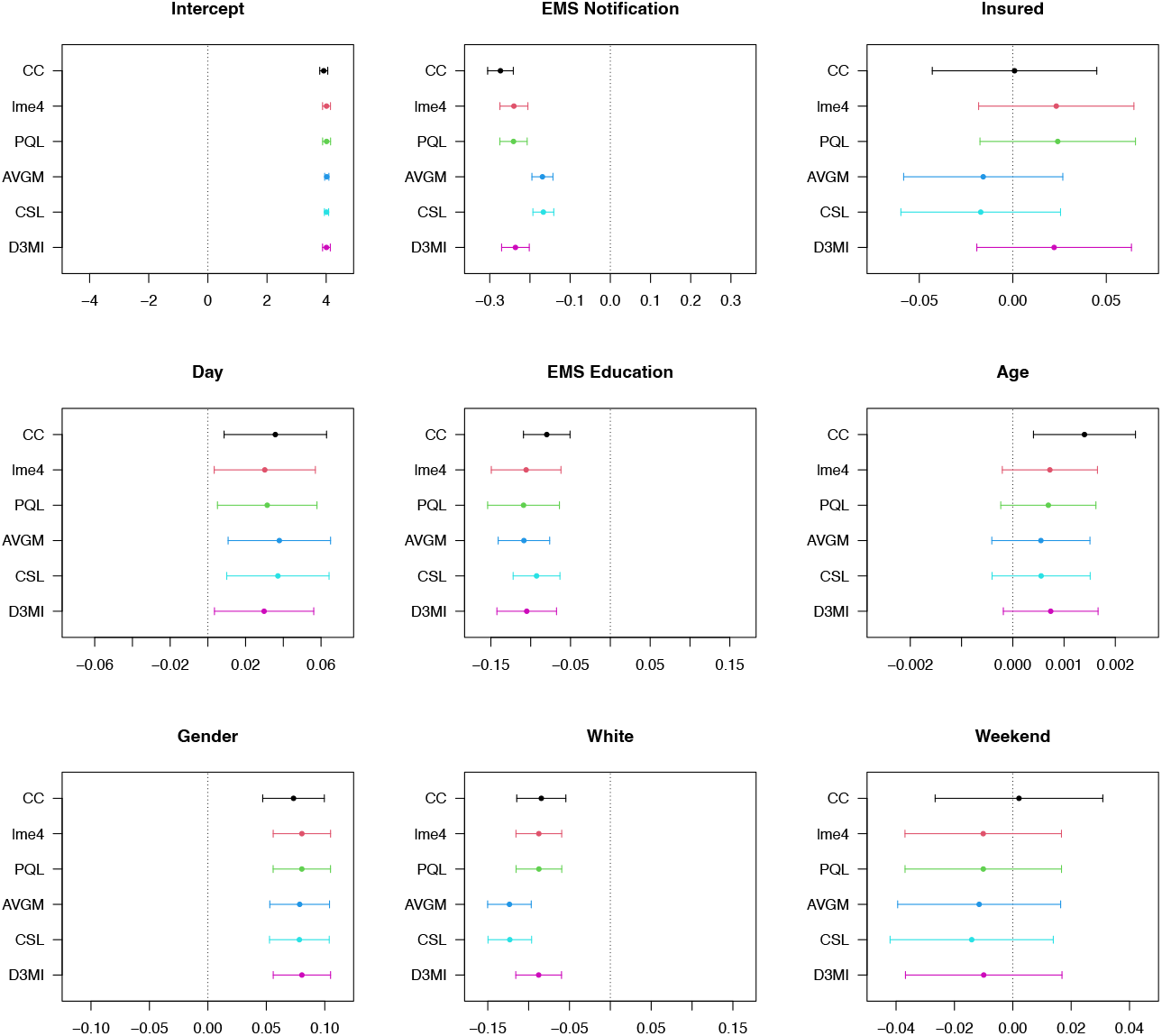
Estimated model parameters using different imputation and analysis models

## DISCUSSIONS

We propose D3MI, a privacy-preserving distributed multiple imputation method that accounts for between-site heterogeneity and within-site correlations. We carefully analyze and evaluate previous work in multilevel multiple imputation and distributed GLMMs. We then propose a practical and effective algorithm that includes specific adaptations and modifications to existing methods. In the design and implementation of D3MI, we prioritize computational and communication efficiency while maintaining satisfactory and consistent performance comparable to the methods using pooled data. The D3MI has several strengths. First, the distributed D3MI enjoys the benefits of centralized GLMM-based multilevel imputation methods, including reduced bias and variability in the estimates, comparing to the current SOTA distributed imputation methods that ignore the clustered structure of the data. Our work further justifies the need for imputation models that can account for the between-site heterogeneity and within-site correlations. Second, D3MI can produce results comparable to those of centralized GLMM-based imputation using “gold standard” GLMM algorithms, demonstrating a successful extension of existing multilevel imputation methods to the distributed setting. Lastly, the computational efficiency of D3MI enables the inclusion of additional random effects (e.g., random slopes) in future research. However, D3MI inherits the limitations of centralized GLMM-based imputation methods and GLMM itself, including the requirement for a healthy number of sites. In addition, these methods do not have a clear advantage over the simpler GLM-based imputation when the between-site heterogeneity is weak. Further research is needed in distributed GLMMs and distributed multilevel imputation to address these limitations.

## CONCLUSION

D3MI is an efficient federated imputation method that accounts for both between-site heterogeneity and within-site correlation. By improving imputation accuracy while preserving privacy, D3MI supports more valid and reproducible analyses in distributed healthcare and other multi-institutional research settings.

## Data Availability

All data produced in the present work are contained in the manuscript.

https://github.com/ly129/D3MI_simulation

## DATA AVAILABILITY

The real-world data analyzed in this article were provided by the Georgia Coverdell Acute Stroke Registry and restrictions apply to the availability of these data. Request for access to the data should be submitted to and approved by the Georgia Coverdell Acute Stroke Registry.

## CODE AVAILABILITY

The D3MI algorithm is implemented in R software and is adapted from the source codes of the R packages `mice` and `pda`.[27, 28] The code is available at https://github.com/ly129/D3MI_simulation/.

## ACKNOWLEDGMENTS

This work was partly supported by the National Institutes of Health, U01-CA274576. The content is solely the responsibility of the authors and does not necessarily represent the official views of the National Institutes of Health. XJ is CPRIT Scholar in Cancer Research (RR180012), and he was supported in part by Christopher Sarofim Family Professorship, UT Stars award, UTHealth startup, the National Institute of Health (NIH) under award number R01AG066749, R01AG066749-03S1, R01LM014520, R01AG082721, R01AG066749, U01AG079847, U01CA274576, and the National Science Foundation (NSF) #2124789.

---

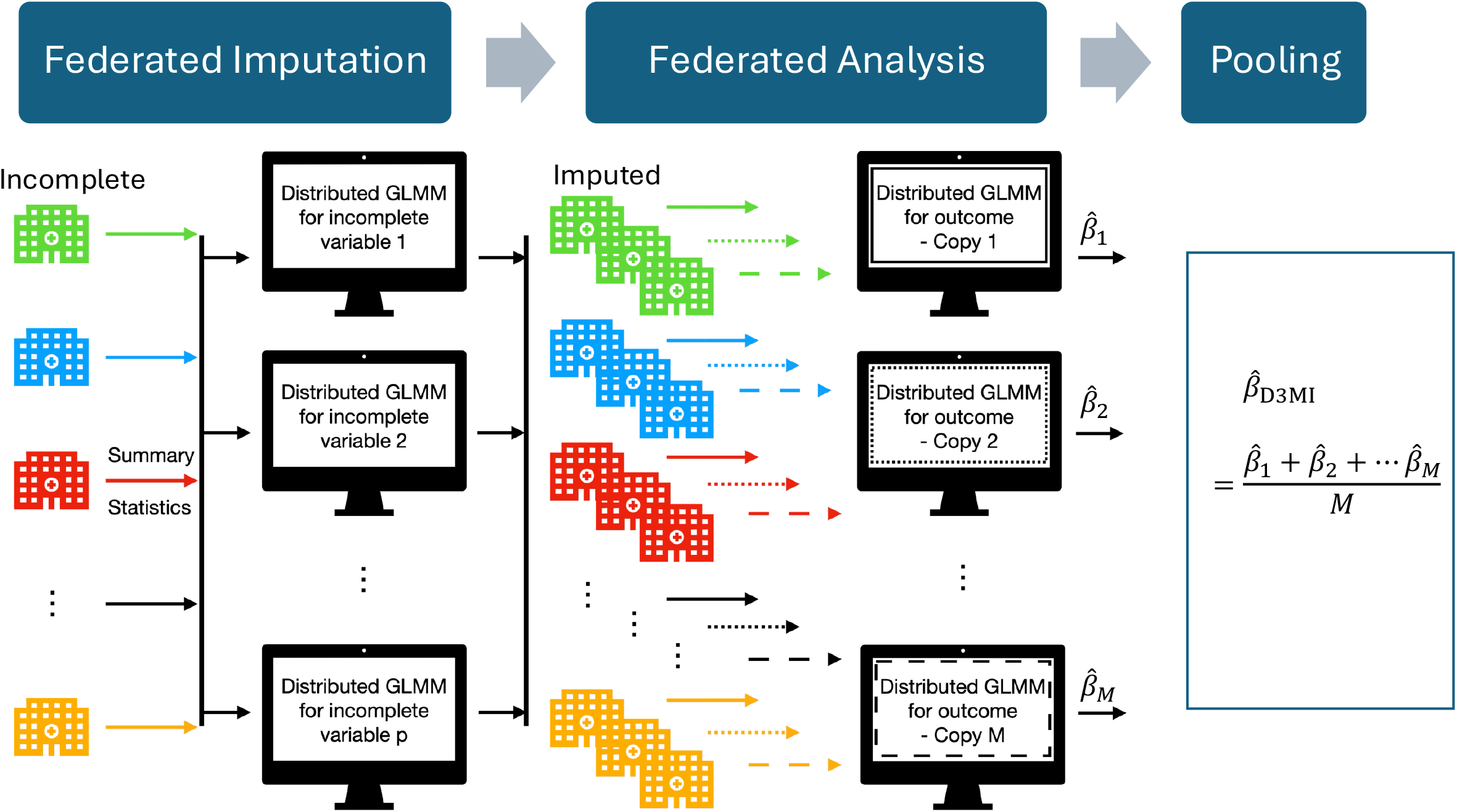

