## Supplementary Material for "D3MI: an efficient and powerful federated imputation method for bias reduction in the analysis of distributed incomplete data by accounting for within-site correlation and between-site heterogeneity"

### **RELATED WORK**

We provide a brief review of existing methods in multilevel multiple imputation for clustered data and computational algorithms associated with GLMM and distributed GLMM.

##### **Multilevel imputation**

Missing data is prevalent in clustered data and appropriate imputation algorithms need to account for the multilevel structure.[1, 2] Under centralized settings, a number of multilevel imputation methods have been proposed and implemented. Existing approaches fall into two categories, namely multilevel imputation by joint modeling and by fully conditional specification.[1] The former works by specifying a single model for all incomplete variables. These imputation algorithms are implemented in the R packages pan and jomo based on Bayesian (generalized) linear mixed models.[3, 4] We refer interested readers to a body of literature on this topic.[1, 3-7] The fully conditional specification approach, on the other hand, iterates univariate multilevel imputation over the variables with missingness. In our case, that is a separate GLMM for each incomplete variable.[8] For each variable with missingness, after a GLMM model is fitted, the next step is to generate random draws of the model parameters. Bayesian Markov chain Monte Carlo (MCMC) method can be used but it can require many iterations of intensive computations for the Markov sequence to converge to generate one draw.[9, 10]. Another option is to leverage the large sample normal approximation of the model parameters,[11] however this method (“2l.norm” in the mice package) only applies to continuous missing variables.[10] An approximate Bayesian approach that requires no iteration for drawing a random set of imputation model parameters has been approached.[9, 10] This method works for both continuous and binary variables with missingness, has been implemented in the mice package in R (method “2l.lmer” and “2l.bin”), and has shown good performance.[8]

##### The computation associated with GLMMs

Consider the GLMM model given in Equation (1) in the main text. The GLMM model parameters of interest are $\left( \beta_{0},\beta,\sigma_{u}^{2} \right)$. Further, the random effect $u_{i}$ in ($1$) for each hospital $i$ can be subsequently estimated through best linear unbiased prediction. The GLMM model likelihood function has the following form,

$$L\left( \beta_{0},\beta,\sigma_{u}^{2} \right)=\prod_{i=1}^{L} \int_{u_{i}} \prod_{j=1}^{n_{i}} f_{y}\left( {y_{ij}|\beta}_{0},\beta,\sigma_{u}^{2} \right)f_{u}\left( u_{i} | \sigma_{u}^{2} \right)du_{i}, (2)$$

where $f_{y}(\cdot)$ is the conditional density of the outcome variable $y$ and $f_{u}(\cdot)$ is the normal density function of the random effect $u$. The likelihood function involves integration over the random effects, which has no closed form unless $f_{y}(\cdot)$ is normal, thus the MLE cannot be obtained using derivative-based optimization algorithms (gradient descent, Newton-Raphson) directly. One option is to numerically approximate the integral using Laplace approximation (LA) or (adaptive) Gaussian-Hermite quadrature (GHQ) and maximize the approximate likelihood.[12-15] The GHQ uses multiple quadrature points to approximate the integral, which lead to accurate approximations at high computational cost. The LA can be viewed as a special case of the GHQ with one quadrature point, which trades off some accuracy for better efficiency. In practice, the R packages lme4, glmmML and glmmTMB use these approximations. These packages adopt different strategies to maximize the approximate likelihood function. lme4[16] primarily uses derivative-free “black-box” optimizers such as the Nelder-Mead algorithm and BOBYQA.[17, 18] The glmmML package[19] and the glmmTMB package[20] use derivative-based optimization with the latter relying on automatic differentiation. Another option is the penalized quasi-likelihood (PQL) approach that uses a first-order Taylor expansion to approximate a GLMM as a linear mixed model (LMM).[21] It is implemented in the R function glmmPQL in the MASS package.[22] The PQL works well when the distribution of the outcome conditional on the random effects is approximately normal.[12] Last but not least, one more option to deal with such integration is to set up an expectation-maximization (EM) algorithm, where the hospital-specific random effects are treated as the latent variable. A Monte Carlo EM algorithm to maximize GLMM likelihood functions has been proposed.[12, 23] and implemented in the MCMCglmm package in R.[24] This algorithm is considered technically challenging and the computation is very expensive [25].

##### **Distributed GLMMs**

Motivated by the restrictions that individual patient data cannot always be shared across hospitals, privacy-preserving distributed GLMM algorithms have been developed. These algorithms are based on different non-distributed GLMM algorithms described in the previous paragraph, hence inherit their respective strengths and limitations. In addition, to yield comparable estimates to the original non-distributed algorithms, the number of communications and the amount of data that need to be transmitted can also be significantly different. We here analyze the computation and communication efficiency of these distributed GLMM algorithms since both are crucial to distributed multiple imputation. First, a distributed version of the Bayesian EM algorithm is proposed.[26] It involves the Metropolis-Hastings algorithm in the E-step and the Newton-Raphson algorithm in the M-step. Both require a round of communication for each update iteration, and the convergence can take up to thousands of iterations for the Metropolis-Hastings algorithm. A second study on distributed GLMM uses a distributed PQL approach (dPQL).[27] The authors show a lossless property of the algorithm, that is, within a few iterations, the dPQL estimates for the GLMM can be almost exactly the same as in the (centralized) PQL estimates for the same model. The communication cost is also relatively low, involving a square matrix and a vector that both have the same dimensionality as the data and a scalar in each iteration. The downside is that the estimates can be inaccurate when the conditional distribution of the outcome variable given the random effects is far from normal. Nonetheless, the PQL approach remains an appealing method for its simplicity and efficiency. Last but not least, a distributed algorithm is developed based on the maximum likelihood approach following the LA and adaptive GHQ approximation.[28] This algorithm is based on the algorithm used by the glmmML package. The algorithm involves evaluating high-order derivatives, which can be numerically unstable. The simulation results show that the algorithm requires up to hundreds of iterations to converge, which means hundreds of rounds of communication, and the estimates have considerable amount of bias. Finally, it is worth mentioning that there has not been a distributed GLMM based on the algorithm used in lme4 that is generally viewed as the go-to package for fitting GLMMs, likely due to the technical difficulty associated with building a distributed derivative-free optimizer.
